# Ensembles of temporal models and forward-backward smoothing for surgical phase recognition

**DOI:** 10.1101/2025.09.02.25334297

**Authors:** Kiran Bhattacharyya, Aneeq Zia, Anthony Jarc

## Abstract

Surgical phase recognition from endoscopic video could enable numerous context-aware technologies that impact efficiency and performance of surgeons and minimally invasive care teams. However, surgical phases can vary greatly (from seconds to minutes) due to patient factors and surgeon workflows along with many other reasons. Given this wide range of phase durations, fixed temporal parameters of neural networks and machine learning models constrain surgical phases with different temporal rhythms. To address this problem, we ensemble neural networks and other machine learning models with different architectures and temporal parameters to recognize surgical phases. The probability estimates of the ensemble are then used as priors for forward-backward smoothing to generate posteriors of surgical phases. We demonstrate the performance of this modeling process on three data sets: 1) robot-assisted inguinal hernia (five phases), 2) robot-assisted training in a porcine model (seven phases), and 3) laparoscopic cholecystectomy (Cholec80, seven phases). The results suggest that this novel method to address varying phases in different procedures holds promise for the future of surgical phase recognition.

## 1 Introduction

Automated detection of critical activities during surgical procedures can advance computer-aided intervention through the development of context-aware applications [26, 10]. Recent work and progress on surgical workflow analysis addresses this problem of detecting surgical phases and workflow from endoscopic video [29, 31, 34, 14].

Moreover, surgery remains to be optimized and surgeons continue to iterate and improve technique and workflows alongside adoption of new enabling technologies. Context-aware applications driven by surgical phase recognition contribute to this surgical innovation as well as to the quantification of its effectiveness [3, 33]. Therefore, developing phase recognition models with the flexibility to accommodate widely varying surgical phases and surgical ontologies [12, 20] is critical in addressing potential bias.

To address this need, we present a modeling process for surgical phase recognition that we evaluated on datasets of different sizes with different statistics of procedure and phase durations. We used an ensemble of deep neural networks [32, 30, 25, 5] and traditional machine learning (ML) pipelines discovered through automated machine learning (AutoML) [23, 24, 19] which were trained with different temporal samples. Furthermore, we used the forward-backward method to smooth the prediction probabilities of the ensemble. We demonstrate the performance of our proposed method on 3 different kinds of minimally-invasive surgical datasets: 1) robot-assisted inguinal hernia—referred to as IHR30, 2) robot-assisted minimally invasive surgery (RMIS) training tasks on a porcine model—RMISTrain150 and 3) Cholec80 [29]. For all datasets, we demonstrate performance in the range of 84–90% for overall accuracy, recall, and precision. We discuss the possible reasons for the robustness of the modeling process across datasets, as well as, how the method can be extended and improved.

## 2 Data and Methods

### Data overview

We evaluate our proposed methodology on three diverse datasets containing 30–150 total cases. IHR30 and Cholec80 had similar distributions of case duration but RMISTrain150 had a much wider range of case durations (Fig. 1). All datasets had phases with short/less-variable durations and long/more-variable durations. However, each had unique distributions and patterns of phase durations (Fig. 1). All datasets had some phases with high variability in their start and stop times (Fig. 1). We provide more details below.

**Fig. 1.**
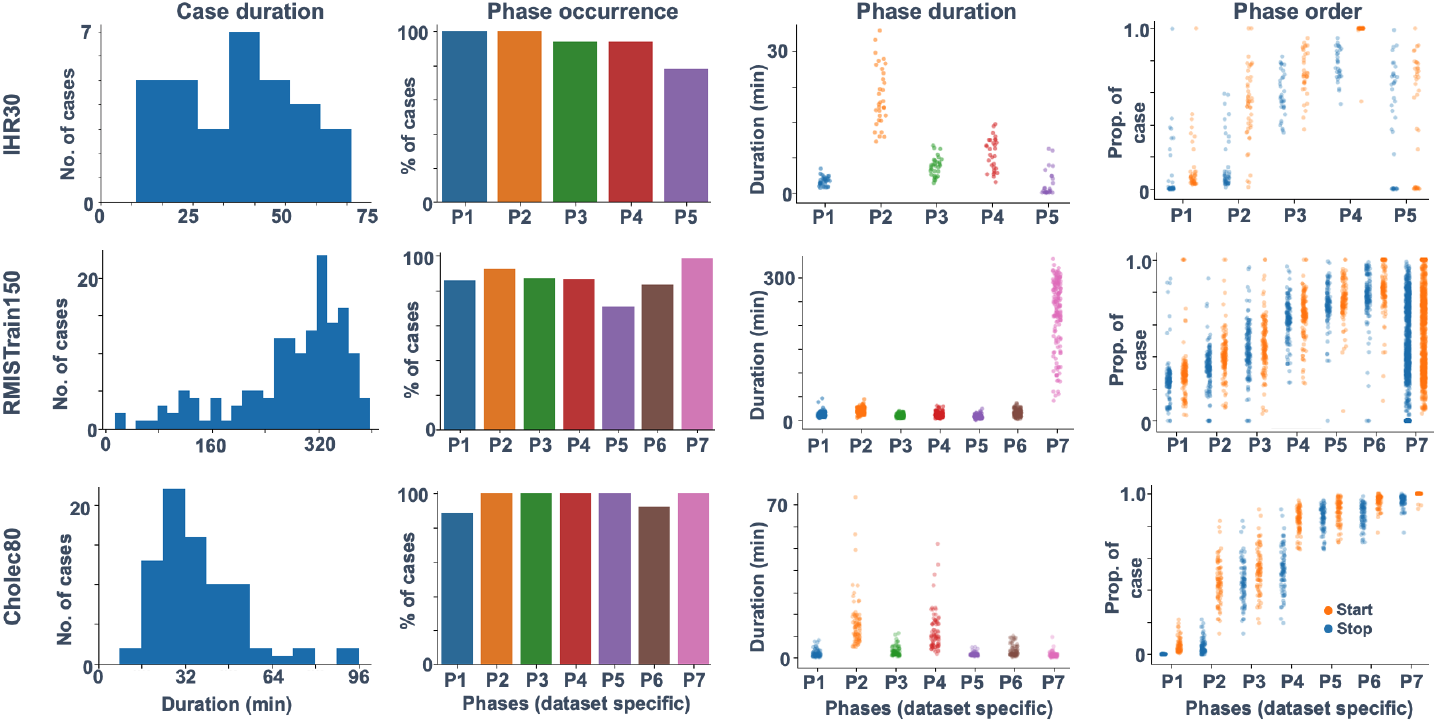
Summary of the datasets used. Each row is a separate dataset and each column is a characteristic of that dataset. For a description of each phase, refer to Data and Methods.

### IHR30 dataset

A set of 30 video recordings of robot-assisted inguinal hernia surgeries collected from the da Vinci Xi system (Intuitive Surgical, Inc.) at 60 fps with a resolution of 720p. Individual surgeries vary in duration from *∼*8–70 minutes (Fig. 1) and contain annotations for the following phases: **P1**: *Creation of peritoneal incision*, **P2**: *Peritoneal flap exploration/reduction of hernia sac*, **P3**: *Mesh placement*, **P4**: *Closure of peritoneum*, and **P5**: *Other/inactivity*.

Most phases occur in every case. However, 2 recordings are missing P3 and P4 since they are not complete and not all cases have P5 (Fig. 1). The phases tend to have an order of appearance in all cases but P5 can occur at many points in the case (Fig. 1).

### RMISTrain150 dataset

Set of video recordings of clinical-like tasks performed by surgical trainees using the da Vinci Xi system on a porcine model. There are 150 total recordings with each lasting *∼*14–400 minutes in duration but surgical phases of interest occur in short periods of time (*∼*2-10 minutes, Fig. 1). The data is collected at 60 fps with a resolution of 720p. The phases in RMISTrain150 are: **P1**: *2-handed suturing*, **P2**: *Uterine horn dissection*, **P3**: *1-handed suturing*, **P4**: *Suspensory ligaments dissection*, **P5**: *Running suture*, **P6**: *Rectal artery/vein dissection*, **P7**: *Other/inactivity*.

Each phase does not necessarily occur in each case (Fig. 1) and sometimes may not be completed. All cases are dominated by long periods of P7. The phases tend to have an order of appearance in each case but P7 occurs at many points in the case and is repeated.

Informed consent was obtained from all individual surgeons and patients whose cases are included in datasets (Western IRB, Inc. Puyallup, WA). Start and stop time annotations for phases in the IHR30 and RMISTrain150 datasets were done manually by subject matter experts. All cases were annotated only once by one annotator with no replicate annotations. The IHR30 and RMIS-Train150 dataset are not publicly available at this time.

### Cholec80

Video recordings of a set of 80 laparoscopic cholecystectomy cases used extensively in surgical phase detection [29]. Each case lasts anywhere from *∼*8-100 minutes (Fig. 1) and contains annotations for the phases: **P1**: *Preparation*, **P2**: *CalotTriangleDissection*, **P3**: *ClippingCutting*, **P4**: *GallbladderDissection*, **P5**: *GallbladderPackaging*, **P6**: *CleaningCoagulation*, **P7**: *GallbladderRetraction*.

### Modeling overview

All datasets were broken into training and testing sets. The train/test split by number of cases for each dataset was the following: IHR*→*30 20/10, RMISTrain*→*150 100/50, and Cholec*→*80 40/40.

We followed a two-step training method using the training set. First, we train a single image-based model which then provides the features for multiple temporal models. All neural networks were trained using categorical crossentropy loss with the Adam optimizer [17]. An early stopping scheme was used to terminate training by monitoring the loss on the validation set—20% of the training set. All models were trained with samples weighted by support as implemented in scikit-learn [4, 16].

Using the training set, we also learned the parameters for the forward-backward algorithm in order to smooth the probability estimates of the model outputs. We ensemble the predictions of all temporal models using soft-voting. All model performances reported are on the test set with and without forward-backward smoothing.

### 2D convolutional neural net (CNN) single-image model

For all datasets, a single-image classifier was trained (Fig. 2A). All videos were down-sampled to 1 fps and resized to (224, 224, 3). The single image classifier had 3 heads used as feature extractors: 1) InceptionResNetV2 [28], 2) ResNet50 [13] and 3) Xception [7]. All three heads had fixed pre-trained weights from the ImageNet dataset [9]. The 2D CNN was trained 20 times with randomly initialized weights of the last two layers and the result with the best performance on the validation set was used for the temporal models.

**Fig. 2.**
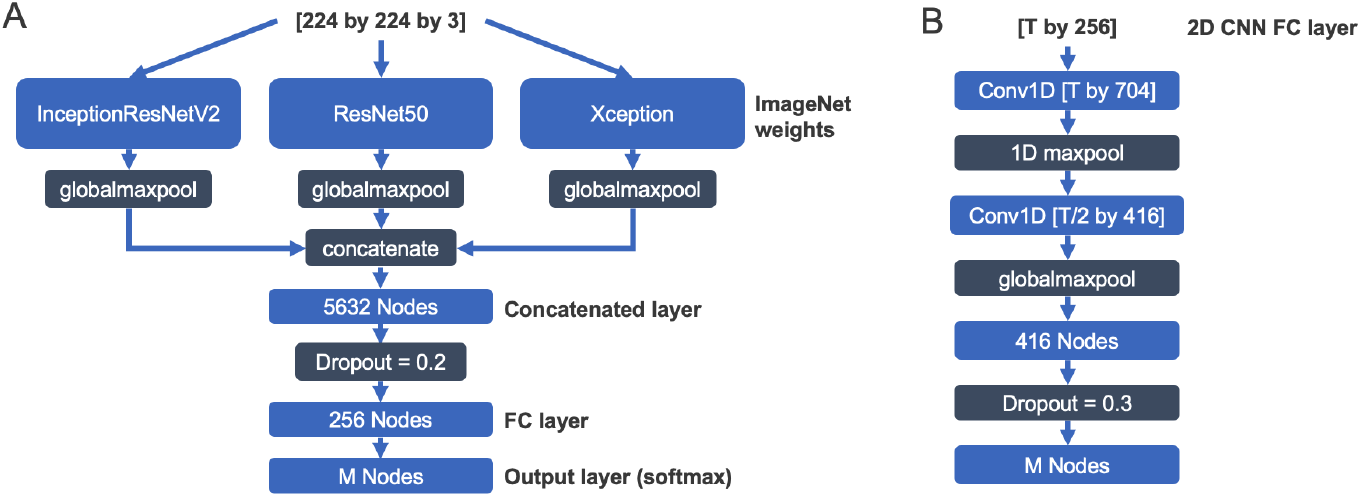
CNN architectures used where *M* (nodes in output) is the number of surgical phases. A) 2D CNN architecture used in this study for single-image model. B) 1D CNN architecture used for temporal models.

### 1D CNN temporal model

The output from the FC Layer of the trained 2D CNN (Fig. 2A) on each frame of the training set was used as features to train 6 different 1D CNN models. For each 1D CNN, a different sliding window of size *T* seconds was used over the FC layer output per-second for all cases in the training set. Successive windows had an overlap of 50% 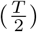 The surgical phase label for each window was the label for the last second of the window. This would produce N samples of *T* by 256 dimensional windows with corresponding N labels. In this case, we tested values of *T* = 8, 16, 32, 64, 96, and 128 seconds. Larger windows reduced N, since the data was sliced into bigger windows. A single 1D CNN was trained for each value of *T*. For the sake simplicity, all 1D CNNs had the same architecture (Fig. 2B) but accepted different input shapes based on the value of *T*.

### AutoML temporal model

The Output layer of the trained 2D CNN (Fig. 2A) on each frame of the training set was used as the features to train 6 different ML pipelines as temporal models (not 1D CNNs). The same sliding windows as described previously were also used to create the training set for these algorithms. However, in this case, the data was flattened to create N samples of *T*×*M* arrays, where *M* is the number of phases for the specific procedure (number of nodes in the Output layer).

For each window of *T*, the architecture for the ML pipeline was discovered with genetic-programming (GP) as implemented in TPOT [24, 19]. TPOT represents ML pipelines as trees of operators (pre-processors, decomposition, feature-selection, and models) where the GP procedure [11] can modify, remove, or insert new sequences of operators into each tree [23]. In each case, the evolutionary process was initialized with a population size of 100 and allowed to run for 20 generations or 10 hours, whichever came first, on 4 parallel processors. The fitness metric for the evolutionary process was balanced accuracy [2].

### Soft-voting ensemble

The predictions of temporal models (1D CNNs and AutoML) were ensembled by taking the argmax of the average of the predicted probabilities. When making inference on the test set with temporal models, the sliding windows were progressed by 1 second increments. Since the temporal models required different windows of time, we ensembled any temporal models able to generate predictions up to the 127^*th*^ second of each case. Starting from the 128^*th*^ second, all temporal models could be ensembled. We ignored the first 7 seconds of all cases.

### Forward-backward smoothing

This algorithm was implemented as presented in Russell and Norvig [27]. The probability estimates for each model or the ensemble were used as the priors. The transition matrix, start probability and stop probability were learned from the training set. To learn these parameters, a case was represented as a succession of surgical phases per-second, [P_1_, P_1_, P_1_,…, P_1_, P_2_, P_2_, P_2_,…]. The transition probability from one surgical phase to the next each second (including auto-transitions) was estimated by counting all transitions in the training set and normalizing by counts. The start and end probability of surgical phases was also estimated by measuring the proportion of times each case in the training set started or ended on a specific phase. Probabilities with values of zero were replaced with a small probability—encountering such an occurrence at least once in the entire training set.

### Software and hardware

All neural networks were implemented in Keras [6] with tensorflow 1.14. All ML algorithms were implemented using scikit-learn [4] and TPOT [19]. All algorithms were trained on a machine with 10 CPU cores, 128 GB of RAM, and 4 RTX 2080 Ti GPUs. However, a 4 CPU cores with 16GB of RAM and one RTX 2080 Ti GPU would be adequate to replicate the work.

All code and models associated with the Cholec80 dataset will be made available upon publication. Code and models associated with IHR30 and RMIS-Train150 cannot be made publicly available at this time.

## 3 Results

### AutoML

The mean balanced accuracy across all model individuals in each generation evaluated on the Cholec80 dataset increased over the first 6–8 generations and then, reached an asymptote (Fig. 3A). Not all runs of AutoML reached 20 generations due to the 10 hour time-limit, but reached at least 8 generations. Since the improvement in mean balanced accuracy over generations were similar for IHR30 and RMISTrain150, those results are excluded to avoid redundancy. Figure 3B shows representative pipelines that were evolved with the AutoML process from temporal models in the Cholec80 dataset where *T* =64 and 128. Pipelines of similar complexity were evolved for other temporal models for Cholec80 and the other datasets. The best evolved pipelines generally had multiple cross-over counts and some mutation counts.

**Fig. 3.**
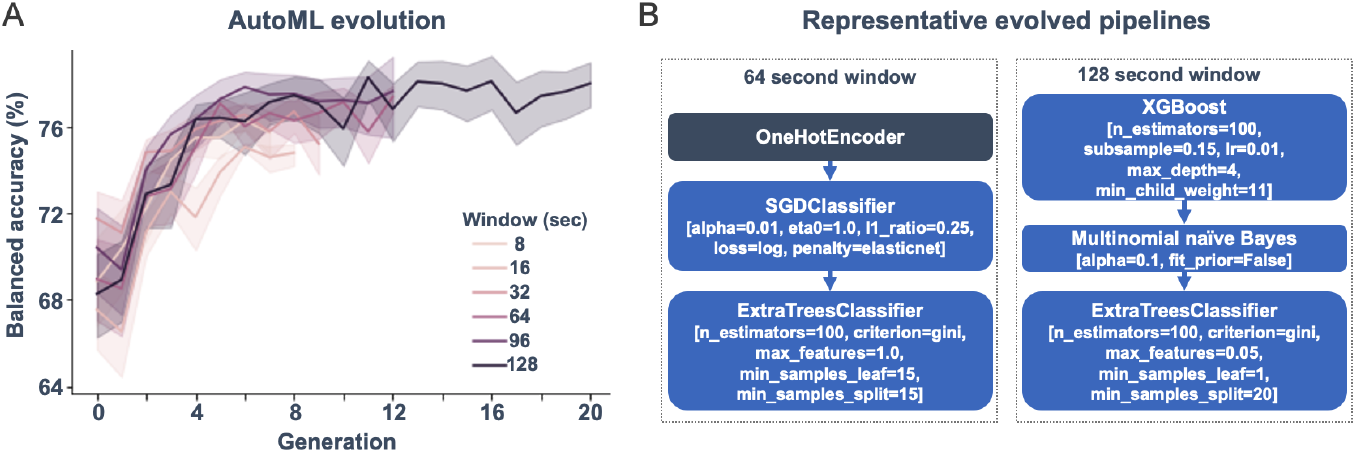
Genetic algorithm based AutoML results from TPOT for Cholec80 dataset. A) Mean balanced accuracy of individuals in each generation of the genetic programming process. B) Representative best evolved pipelines for *T* =64 and *T* =128 second windows.

### IHR30

The single frame model performs quite well in this dataset achieving nearly 83% accuracy after smoothing (Fig. 4A). Interestingly, temporal models where *T* was 8–32 seconds performed better after smoothing than models with longer *T*. Critically, the ensemble of all temporal models performed better than any single one. The ensemble model was best at identifying phases P2 and P3 (Fig. 4B). This may be because P2 tended to be longest phase (Fig. 1) and P3: *Mesh placement* had the salient visual features of a hernia mesh. The ensemble model struggled most to identify P5 possibly because this phase was uncommon.

**Fig. 4.**
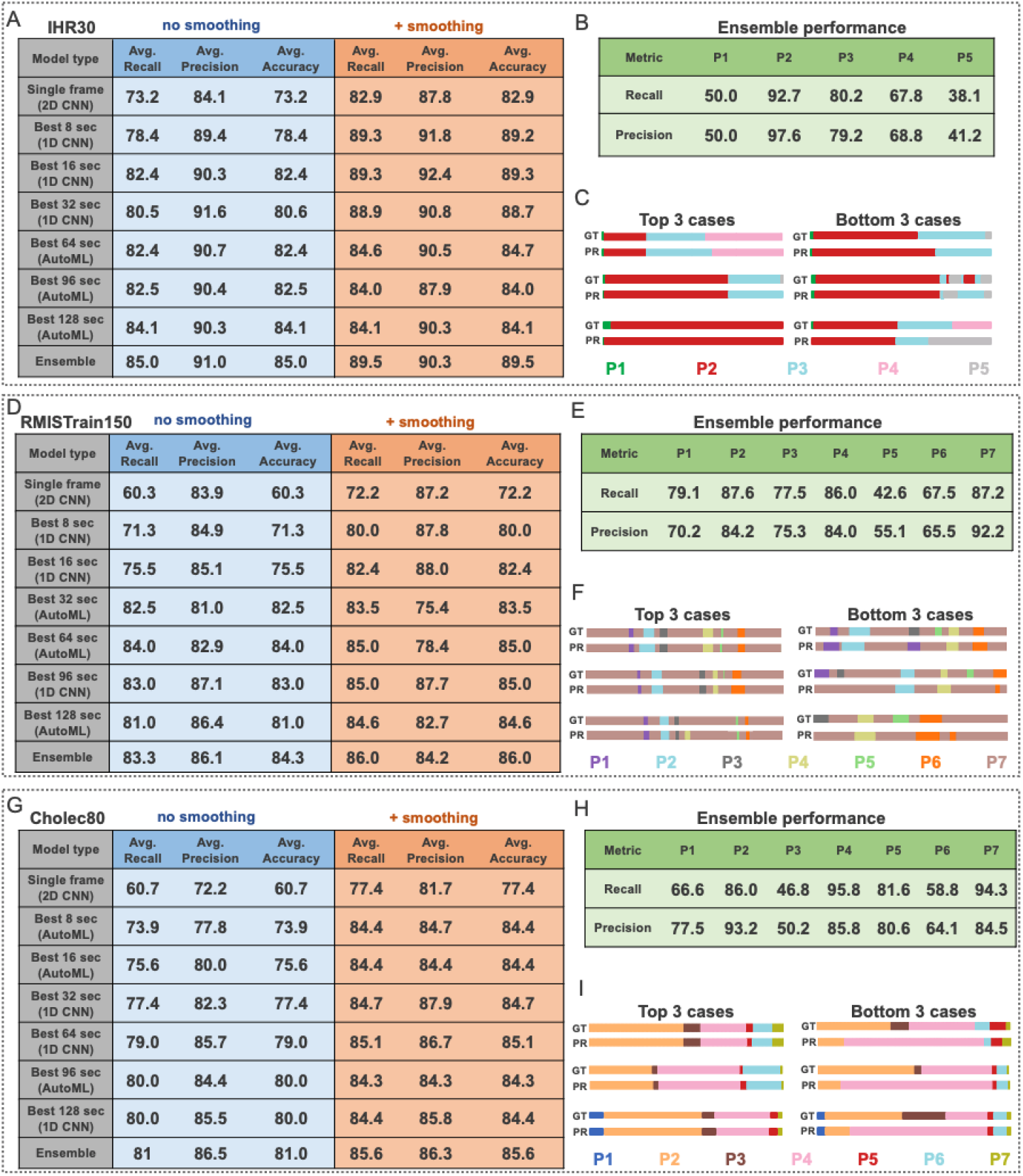
(A–C) IHR30 results. (D–F) RMISTrain150 results. (G–I) Cholec80 results. A, D, and G) Weighted-average recall, weighted-average precision, and accuracy on the testing set averaged for all cases. The table only includes the temporal model (1D CNN or AutoML) for each time window with the highest accuracy. B, E, and H) Performance of the ensemble on each surgical phase. C, F and I) Ground truth (GT) and predictions (PR) of the ensemble model for cases with the best and worst performance.

### RMISTrain150

Similar to IHR30, temporal models with specific values of *T* had higher accuracy than others (Fig. 4D, 64 and 96 sec). Smoothing provides consistent improvement for all models and, again, the ensemble model outperforms any single model. The ensemble has the best performance (80–92% recall/precision) for P2, P4, and P7 (Fig. 4E). However, the model has the lowest performance on P5 possibly since this was sometimes confused with P7 (Fig. 4F). Regardless, the ensemble model is able to successfully identify short phases embedded in long stretches of P7 (Fig. 4F).

### Cholec80

Interestingly, while temporal models increase in accuracy with increasing *T* before smoothing, all temporal models have comparable accuracy after smoothing (Fig. 4G), with *T* = 64 seconds slightly outperforming others. Nonetheless, the ensemble model performs better than any single model and achieves performance comparable to existing benchmarks [29, 14, 15, 1, 8]. Importantly, the ensemble model had 80–95% recall/precision for P2, P4, P5, and P7 which are a combination of longest and shortest phases. However, the model struggled the most with P3 and confused it with P4 which immediately followed in all cases (Fig. 4H and I). This may be because the surgeon is interacting with similar anatomy during these phases.

## 4 Discussion

We demonstrate the performance of a novel modeling process for surgical phase recognition that supports clinically-relevant variability around temporal rhythms without sacrificing accuracy. In particular, we examine the approach on distinct datasets with different number of samples, phases, and statistics of phase duration and sequence. We make three important observations of our results. Firstly, our results approximate current state-of-the-art for Cholec80. Secondly, the modeling approach performs well on IHR30 where there were only 20 cases in the training set. Finally, the approach produces good performance on RMISTrain150 which has highly variable case duration, phase duration, and phase start/stop times.

The robustness across varied datasets suggests this approach could support rapid generalizability of phase recognition to new procedures and corresponding datasets with unique characteristics. We believe that the strength of this approach could be due to 1) the combination of multiple temporal models with varying parameters and 2) the forward-backward smoothing which imposes temporal patterns over the entire case through belief propagation.

There exist several areas for improvement. First, there may be an interaction between dataset size and procedure type complexity which can influence our results, and this needs to be explored further. Moreover, each aspect of the process can be further improved. The feature extraction heads of the 2D CNN can be fine-tuned by pre-training on all datasets with a self-supervised task [22]. Training-free neural architecture search [21] can be implemented to discover 1D CNN designs specific to each value of *T*. Finally, model-based ensembles [18] of all temporal models could improve upon the soft-voting strategy used here.

Due to the robust performance on varying datasets and the methods of improvement described, we believe this process could contribute a novel and promising approach to address surgical phase recognition in real-world and continually evolving surgical environments in order to enable improved surgeon performance.

## Data Availability

Two of the datasets used in this study are available by request; the Cholec80 dataset and the RMISTrain150 dataset. Please note that the RMISTrain150 dataset is now referred to as the SurgVU dataset released recently. One other dataset used in this study is not yet publicly available but the authors are working towards this goal and hope to release it soon.

https://camma.unistra.fr/datasets/

https://arxiv.org/abs/2501.09209

## Notes

### Competing Interest Statement

Authors are employees at Intuitive Surgical, Inc.

### Funding Statement

This study did not receive any funding.

### Author Declarations

Ethics approval was provided for this work by Western IRB, Inc. Puyallup, WA. Now known as WCG Clinical Services.

